# Long-Term Exposure to Outdoor Air Pollution and COVID-19 Mortality: an ecological analysis in England

**DOI:** 10.1101/2020.08.13.20174227

**Authors:** Zhiqiang Feng, Mark Cherrie, Chris Dibben

## Abstract

There is an urgent need to examine what individual and environmental risk factors are associated with COVID-19 mortality. This objective of this study is to investigate the association between long term exposure to air pollution and COVID-19 mortality. We conducted a nationwide, ecological study using zero-inflated negative binomial models to estimate the association between long term (2014-2018) small area level exposure to NO_x_, PM_2.5_, PM_10_ and SO_2_ and COVID-19 mortality rates in England adjusting for socioeconomic factors and infection exposure. We found that all four pollutant concentrations were positively associated with COVID-19 mortality. The increase in mortality risk ratio per inter quarter range increase was for PM_2.5_:11%, 95%CIs 6%-17%), PM_10_ (5%; 95%CIs 1%-11%), NOx (11%, 95%CIs 6%-15%) and SO_2_ (7%, 95%CIs 3%-11%) were respectively in adjusted models. Public health intervention may need to protect people who are in highly polluted areas from COVID-19 infections.

## Introduction

It is well established that long term exposure to air pollution is harmful to human health and has led to excess morbidity and mortality (Hoek et al 2013; Brunekreef and Holgate, 2002; Samet et al 2000). The mechanism points to both direct and indirect systemic impact on the human body through increasing oxidative stress and inflammation, resulting in respiratory, cardiovascular, and immune system dysfunction and deterioration in circulation (Brunekreef. and Holgate, 2002; Brook et al 2010; WHO 2004; Zanobetti and Schwartz, 2009).

The COVID-19 pandemic that is caused by a novel coronavirus (SARs-COV-2), has, as of July, 2020 claimed over half a million lives worldwide and also resulted in serious socio and economic consequences. Since the December of 2019 when the pandemic started medical and epidemiological studies have improved our knowledge on the aetiology of COVID-19-related disease. However, there is still considerable unknown on its pathogenesis, as well as factors contributing to disease severity. Much attention has been paid to individual factors. Research worldwide has showed many individual risk factors that are associated with the risk of COVID-19 related death including older age, underlying health conditions, belonging to an ethnic minority group, and lower socioeconomic status (Verity et al 2020; Travaglio et al 2020, Williamson et al, 2020). Environmental factors, such as air pollution, may play an important role in increasing susceptibility to severe outcomes of COVID-19. But their impacts have been understudied so far.

In most cases, COVID-19 results in mild symptoms. However, it may lead to an excessive inflammatory response causing Acute Respiratory Distress Syndrome (ARDS) and death (Guan et al, 2020; Cao, 2020). There is clear overlap between the symptoms of Covid-19-induced ARDS and long term exposure to air pollution; it is highly likely that the underlying mechanism operates similarly (Qu et al 2020; Cole et al 2020). Air pollutants can increase the risk of COVID-19 death in different ways. Major pollutants such as fine particulate matter (PM2.5, PM10), nitrogen oxide (NOx), andsulphur dioxide (SO_2_), have been suggested to be associated with severity of COVID-19 conditions. This is similar to the pathways that they affect respiratory and cardiovascular mortality. For example, NO_x_ can reduce lung activity and increase infection in the airway. SO_2_ has significant impacts on TNF, 1 IL 2 1 b, IL-6, IL-8, IL-17 and IL-18, they have a prominent role in inflammation of the respiratory and systemic systems, thus increasing the risk of developing coronavirus. On the other hand, PM10/2.5 by penetrating the depth of lung not only can paralyzes the ciliums airway, but also lead to chronic respiratory tract inflammation (Comunian et al 2020; Liang et al 2020).

Previous findings on the severe acute respiratory syndrome (SARS) have shown that exposure to pollution was linked to the high mortality related to the Sars-Cov-1 virus (Cui et al., 2003) with a positive association being observed between poor air quality and high SARS case-fatality rate in the Chinese population. A limited number of studies have explored the association of air pollution with COVID-19 related mortality (Wu et al 2020; Liang et al 2020; Ogen 2020; Conticini et al 2020). The conclusions are so far mixed with some identifying positive associations while others not.

In this study we explore the association between air pollution and COVID-19 mortality across a large number of small area data in England. We used ecological data analysis based on middle super output areas and focused on four key air pollutants, particulate matter 2.5, particulate matter 10, nitrogen oxide and sulphur dioxide.

## Data

The COVID-19 death data is acquired from ONS (Office for National Statistics). (https://www.ons.gov.uk/peoplepopulationandcommunity/birthsdeathsandmarriages/deaths/bulletins/deathsinvolvingcovid19bylocalareasanddeprivation/). Data include all deaths of three months from 1^st^ of March to the 31st of May, 2020 when the majority of deaths in England occurred. COVID-19 death referred to a death that COVID-19 was mentioned in the death certificate, with a delay of usually five days between occurrence and registration. The data were the number of cumulative deaths in Middle Lower Super Output Areas (MSOA, average population was 8200) in England.

The age composition and population density data was from the ONS mid-year population estimates in 2018 (https://www.ons.gov.uk/peoplepopulationandcommunity/populationandmigration/populationestimates/datasets/lowersuperoutputareamidyearpopulationestimates). The ethnicity, care home, and commuting data were from the 2011 census. Income deprivation data were from the index of multiple deprivation (IMD) statistics of 2019, provided by the Ministry of Housing, Communities & Local Government (MHCLG, 2019).

We measured the percentage of the population from a minority group in each MSOA as the proportion that reported their ethnic group as Black, South Asian, or Chinese in the 2011 Census. The percentage of people who reported to have limiting long term illness was also derived from the 2011 census. Our measure of deprivation was the income deprivation domain from the IMD 2019 score. The income deprivation score measures the percentage of the population experiencing deprivation relating to low income, based on a non-overlapping count of people receiving welfare benefits for low-income. It includes those who are out-of-work and those who are in work but have low earnings (MHCLG, 2019). Population estimate data provided by the ONS were used to calculate percentages of the population aged 65 and over for MSOAs.

In order to adjust for viral exposure we include measures including population density, means of transport for commuting and the level of viral transmission by days since the day when there first was 10 confirmed COVID-19 case in the local authority. Population density was computed using the mid-year population estimates in 2018. The Public Health England (PHE) data (https://coronavirus.data.gov.uk/) was used to calculate the number of days since the day when 10 laboratory confirmed cases in a local authority were identified. The 2011 census data on percentage of residents using different forms of transport for commuting (bus, train, tube) were also derived to measure virus spread in a local area.

Four pollutants were explored in this research: PM_2.5_, PM_10_, NO_x_ and SO_2_. They were obtained from DEFRA (Department for Environment, Food and Rural Affairs) modelled annual average pollutant concentrations at a 1km by 1km resolution from 2014 to 2018. The five year average of four pollutant concentrations were used to indicate long term exposure to air pollution. Concentration values for each of the pollutants were for 1km grid-squares (https://uk-air.defra.gov.uk/data/pcm-data), which was interpolated to MSOA level using the point-in-polygon method. For middle-layer Super Output Areas that did not have grid points falling within them, data from the nearest point of the air quality grid was assigned. These four pollutants have been shown to be associated with all cause mortality, respiratory mortality and cardiovascular mortality in England (Carey et al 2013) and have been explored in research on COVID-19 mortality (Cole et al 2020; Wu et al 2020; Liang et al 2020).

## Methods

We estimated the relationship between air pollution and COVID-19 deaths using a zero-inflated negative binomial mixed model (ZINB) (Venable & Ripley, 2002; Zhang et al 2017). We tested the zero-inflation assumption and the result showed that there were substantially more zeros in the observed outcome compared than expected from a Negative Binomial process. These zeros arise because the absence of COVID-19 cases (and presumably the SARS-CoV-2 virus) in some MSOAs on/before the end of May, 2020 making them ineligible to experience a COVID-19 death.

The number of COVID-19 deaths was the outcome and the logged population size was the offset. There are two sub-models in the ZINB model. The first is a count sub-model that estimates the association between COVID-19 deaths and pollutants. The second is a zero sub-model that takes care of the excess of zeros that may be produced by MSOAs not yet eligible for COVID-19 deaths (e.g., due to the absence of confirmed COVID-19 cases) and unlikely to have COVID-19 deaths as of 31st May, 2020. We also included a random intercept by lower tier local authority (N=315) to account for potential correlation in MSOAs within the same local authority, due to similar socio-cultural, behavioural, and healthcare system features. All confounders were normalised before entering the model. Three sets of models were fitted separately. The first set of models were adjusted at the MSOA level for: percentage of residents: aged 65+, Black, South Asian, Chinese, in care homes. Population density and the number of days from the day when 10 infection cases were identified were also adjusted. For the zero inflation model, in addition to socioeconomic variables mentioned above percentages of public transport commuters and particular matters (PM_2.5_) in 2018 were adjusted for. The second and the third sets of models expanded the first set by including percentage of low income people and percentage of people with limiting long term illness respectively to the first set of models.

hEffect estimates were presented as mortality rate ratios (MRR) with 95% confidence intervals. The MRR can be interpreted as the relative increase in the COVID-19 death rate associated with a 1 unit increase in long-term average pollutant concentrations among MSOAs eligible to experience a COVID-19 death. We reported the mortality rate ratio per inter-quartile range increase for pollutant concentrations. Statistical analysis was conducted in R. Data and syntax are available upon request.

## Results

The total number of COVID-19 deaths were 44,359 in the three months from March to May with an average of 7 deaths across MSOAs. The total number of MSOAs included in our main analysis is 6,971, of which 237 (3%) had not reported any COVID-19 deaths by the end of May, 2020. The high death rates occurred mostly in London, Liverpool, Sunderland, Sheffield, Birmingham, and Cumbria (Figure 1). MSOAs within Greater London had the highest concentrations for PM_10_, PM_2.5_, and NOx. By contrast, the highest concentrations for SO_2_ existed in Northwest, and Yorkshire and the Humber. The average annual concentrations of four pollutants are correlated with each other. PM_10_, PM_2.5_, NOx have strong correlations between them (PM_2.5_ vs PM_10_ 0.97; PM_2.5_ vs NOx 0.75; PM_10_ vs NOx 0.71). In contrast, these three pollutants have low-moderate correlation with SO_2_ (PM_2.5_ vs SO_2_: 0.13, PM_10_ v SO_2_: 0.09; NOx vs SO_2_: 0.41).

**Figure 1.**
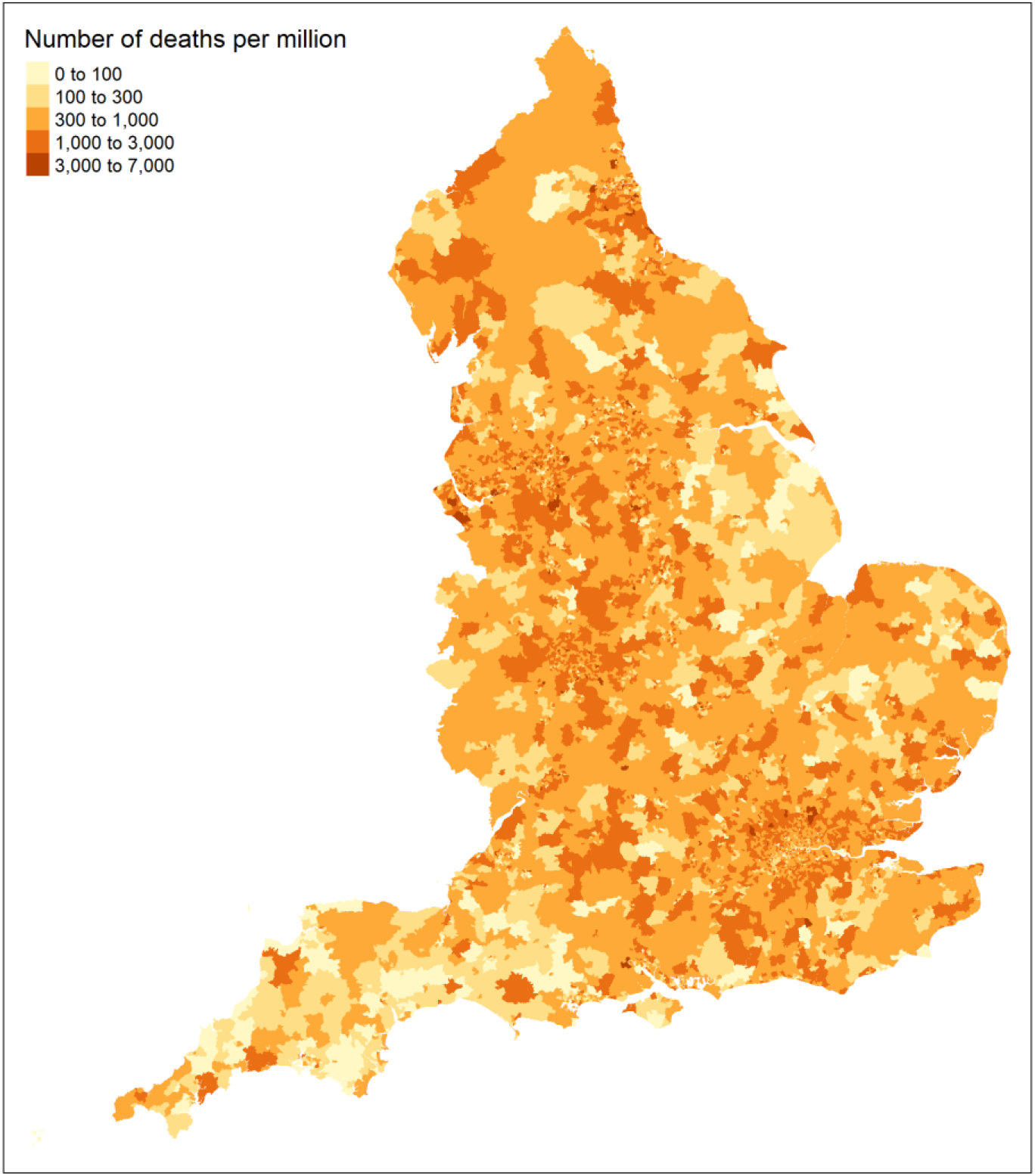
COVID-19 deaths (March to May) per million in middle super output areas.

There was considerable variation in terms of demographic structure, population density, travel behaviour, low income and prevalence of limiting long term illness. For examples, people aged 65 and over ranged from 1% to 52%. Percentage of black people in a local middle super output area varied from no black people at all to over half of the local population. Days from 10 confirmed cases were also markedly different in local authorities from 48 days to 88 days during the observation period (Table 1).

**Table 1.**
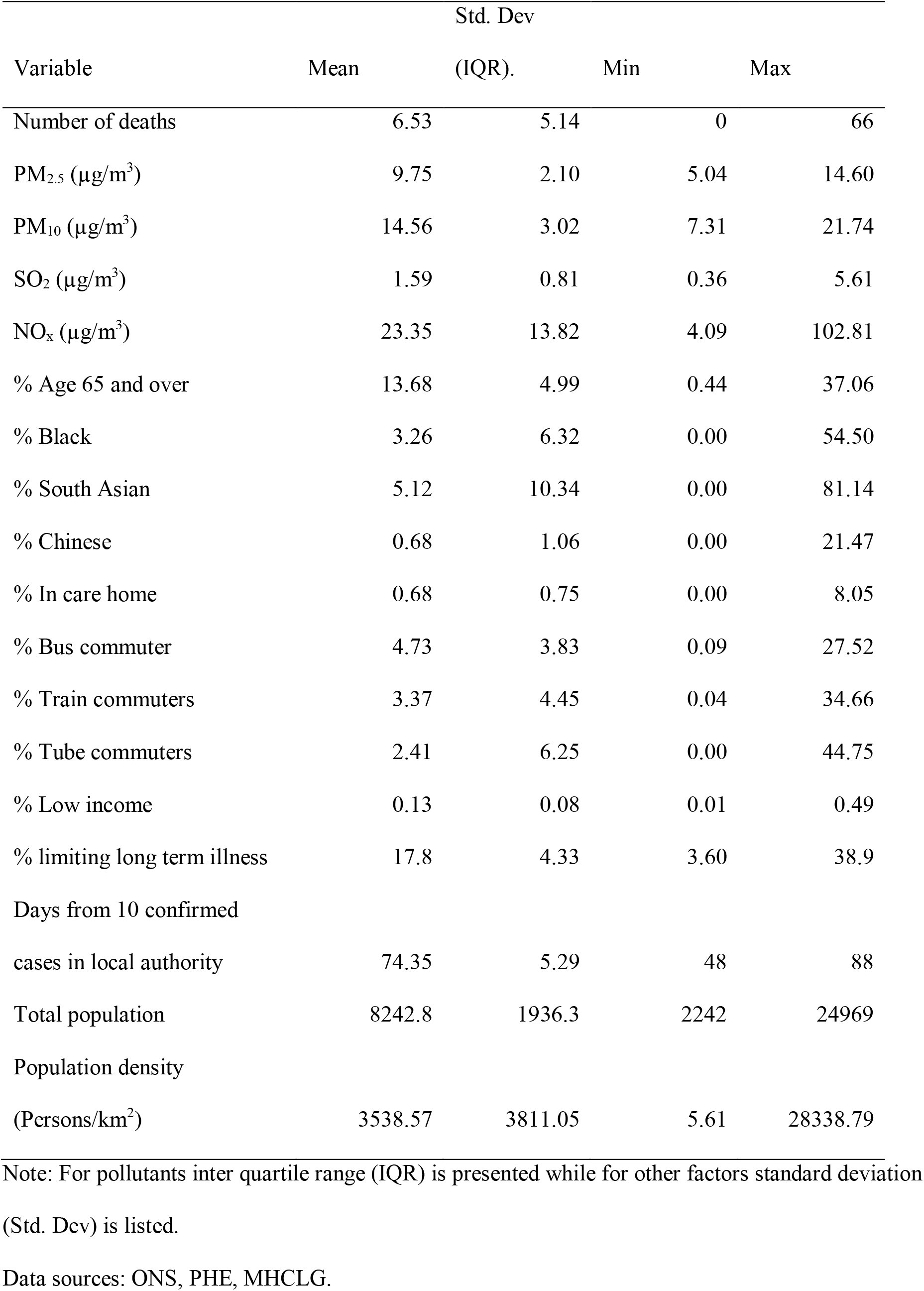
Summary statistics of variables.

The results from the models are shown in Table 2. Model 1 refers to the model that adjusted for socioeconomic variables while Model 2 added the variable of low income and model 3 added the limiting long term illness variable. We reported the association with socioeconomic and infection spread variables first (Model 1). As expected areas with a higher proportion of people aged over 65, higher proportion of Black and South Asian, higher proportion of people in care home, higher level of low income people were associated with higher risks of COVID-19 deaths (Model 2). In Model 3, percentage of limiting long term illness was found to be associated with higher risks of COVID-19 deaths. Areas where there was a longer duration in coronavirus infection was also linked to a higher risk of COVID-19 deaths. In contrast, areas with a higher proportion of Chinese were related to lower COVID-19 mortality risk. Population density was not a significant predictor of COVID-19 death.

Associations with COVID-19 deaths were positive for all four pollutants. Based on the first set of models (Model 1), after adjustment for age composition, population density, ethnicity, and days since 10 confirmed cases in the local authority mortality rate ratios relating to PM_2.5_, PM_10_, NO_x_ and SO_2_ per inter-quartile range increase were separately 1.10 (95%CI 1.04-1.16); 1.04 (95%CI 0.99-1.09); 1.13 (95%CI 1.08-1.18); 1.12 (95%CI 1.08-1.16). An increase of inter quartile range (2.1 μg/m3) in PM_2.5_ was associated with a 10% increase in the risk of COVID-19 mortality independent of a number of socioeconomic risk factors. For PM_10_, NO_x_ and SO_2_, the equivalent increases in MRR were respectively 4%, 13% and 12%. However, the association with PM_10_ was not significant. When the additional low income variable and additional limiting long term illness variables were separately adjusted for the positive association with the mortality risk of these four pollutants remained largely unchanged for PM_2.5_, NO_x_ and SO_2_. However, the association with PM_10_ changed to be significant. The association with SO_2_ was attenuated down to 1.09 (Model 2, 95% CI, 1.05-1.12).

**Table 2.** Estimated mortality rate ratios from the zero-inflated negative binomial mixed models.

| Variable | Model | Model 2 | Model 3 |
| --- | --- | --- | --- |
| PM<sub>2.5</sub> | 1.10(1.04-1.16) | 1.12(1.06-1.17) | 1.11(1.06-1.17) |
| Population density (persons/Km<sup>2</sup>) | 0.99(0.97-1.02) | 0.98(0.95-1.01) | 0.98(0.95-1.01) |
| % Age 65 and above | 1.16(1.13-1.19) | 1.23(1.19-1.26) | 1.13(1.1-1.16) |
| % Black | 1.07(1.04-1.09) | 1.03(1.00-1.06) | 1.05(1.02-1.07) |
| % South Asian | 1.04(1.02-1.06) | 1.03(1.01-1.05) | 1.04(1.02-1.06) |
| % Chinese | 0.92(0.9-0.93) | 0.94(0.92-0.96) | 0.94(0.92-0.96) |
| % Care home | 1.28(1.26-1.3) | 1.27(1.25-1.29) | 1.25(1.24-1.27) |
| Days since 10 confirmed cases | 1.20(1.16-1.26) | 1.23(1.18-1.28) | 1.23(1.18-1.28) |
| % Low income |  | 1.11(1.08-1.13) |  |
| % Limiting long term illness |  |  | 1.14(1.11-1.16) |
| PM<sub>10</sub> | 1.04(0.99-1.09) | 1.06(1.01-1.11) | 1.05(1.01-1.10) |
| Population density (persons/Km<sup>2</sup>) | 1.01(0.98-1.03) | 0.99(0.97-1.02) | 0.99(0.96-1.02) |
| % age 65 and above | 1.16(1.13-1.18) | 1.22(1.19-1.25) | 1.12(1.1-1.15) |
| % Black | 1.07(1.04-1.1) | 1.03(1.01-1.06) | 1.05(1.02-1.08) |
| % South Asian | 1.04(1.02-1.06) | 1.03(1.01-1.05) | 1.04(1.02-1.06) |
| % Chinese | 0.92(0.9-0.93) | 0.94(0.92-0.96) | 0.94(0.92-0.96) |
| % care home | 1.28(1.26-1.3) | 1.27(1.25-1.29) | 1.25(1.24-1.27) |
| Days since 10 confirmed cases | 1.22(1.17-1.27) | 1.24(1.19-1.29) | 1.24(1.19-1.3) |
| % Low income |  | 1.11(1.08-1.13) |  |
| % Limiting long term illness |  |  | 1.14(1.11-1.16) |
| NO<sub>x</sub> | 1.13(1.08-1.18) | 1.11(1.07-1.16) | 1.11(1.06-1.15) |
| population density | 0.99(0.96-1.02) | 0.98(0.95-1.01) | 0.98(0.95-1.01) |
| % age 65 and above | 1.16(1.14-1.19) | 1.22(1.19-1.26) | 1.13(1.11-1.16) |
| % Black | 1.06(1.04-1.09) | 1.03(1.01-1.06) | 1.05(1.02-1.07) |
| % South Asian | 1.03(1.01-1.05) | 1.03(1.01-1.05) | 1.04(1.02-1.06) |
| % Chinese | 0.90(0.89-0.92) | 0.93(0.91-0.95) | 0.93(0.91-0.95) |
| % care home | 1.28(1.26-1.29) | 1.27(1.25-1.29) | 1.25(1.23-1.27) |
| Days since 10 confirmed cases | 1.18(1.13-1.23) | 1.21(1.16-1.26) | 1.22(1.17-1.27) |
| % Low income |  | 1.10(1.08-1.12) |  |
| % Limiting long term illness |  |  | 1.13(1.10-1.15) |
| SO<sub>2</sub> | 1.12(1.08-1.16) | 1.09(1.05-1.12) | 1.07(1.03-1.11) |
| Population density | 1.00(0.97-1.03) | 0.99(0.97-1.02) | 0.99(0.96-1.02) |
| % age 65 and above | 1.18(1.15-1.21) | 1.22(1.19-1.26) | 1.14(1.11-1.17) |
| % Black | 1.07(1.05-1.1) | 1.04(1.02-1.07) | 1.06(1.03-1.08) |
| % South Asian | 1.04(1.02-1.06) | 1.03(1.01-1.05) | 1.04(1.02-1.06) |
| % Chinese | 0.92(0.9-0.94) | 0.94(0.92-0.96) | 0.94(0.92-0.96) |
| % care home | 1.28(1.26-1.29) | 1.27(1.25-1.29) | 1.25(1.24-1.27) |
| Days since 10 confirmed cases | 1.22(1.17-1.27) | 1.24(1.19-1.3) | 1.25(1.20-1.30) |
| % Low income |  | 1.09(1.07-1.12) |  |
| % Limiting long term illness |  |  | 1.12(1.10-1.15) |

Note: 95% confidence intervals are presented in bracket
Data sources: ONS, PHE, MHCLG.

## Discussion

Using data from various sources including small area COVID-19 death registrations, census, income deprivation and COVID-19 cases from public health bodies of England this study examined the association between ambient air pollution and COVID-19 mortality. We found statistically significant associations between four pollutants risk of COVID-19 mortality. These associations were independent of demographic, socioeconomic factors and viral exposure.

The larger effect was observed for PM_2.5_ and NO_x_ than that for PM_10_ and SO_2_ based on the model that included both percentage of low income or limiting long term illness with an increase of inter-quartile range leading to 12% and 11% increase in mortality rate ratio. The effect of PM_10_ and SO_2_ was similar with an increase of inter-quartile range leading to 6% and 8% increase in mortality rate ratio of COVID-19 death.

The results of the study suggest that long term exposure to air pollution is associated with the most severe COVID-19 outcomes. These findings are in line with previous studies that showed association with respiratory and cardiovascular mortality (Carey et al 2003). They are also consistent with findings that air pollution exposure increased the risk of death during the severe acute respiratory syndrome (SARS) outbreak in 2003 (Cui et al 2003). They provided further support to previous studies that observed 78% of the COVID-19 deaths occurred in the most polluted five regions with the highest NO2 levels (Ogen, 2020) and another study that found a correlation between high levels of air pollution and high death rates in Italy (Conticini et al, 2020). Nevertheless these two studies had severe limitations because of their methodologies. The paper by Ogen (2020) used two month pollution data as long term exposure measures and both failed to adjust for potential confounders. Travaglio et al. (2020) took a similar approach but control only for differences in population density and across only 7 relatively large regions. Studies based on U.S. were rigorous in terms of the data and methodology. However, their finding have been mixed. For example, while some found a positive association with PM2.5 (Wu et al 2020) others did not (Liang et al 2020; Knittel and Ozaltun, 2020). In the work by Liang et al (2020) they examined three NO2, PM2.5 and O3 and found that only NO2 showed consistent positive association with COVID-19 mortality.

The strengths of this analysis include air pollution measures based on well validated approaches used in a large number of previous studies (Carey et al 2013; Dibben and Clemens, 2015). Our analysis utilised COVID-19 death data up to 31^st^ May 2020 allowing us to capture almost the entire course of the first phase of the pandemic in England and hence much more fully than the previous studies which have examined data up to only March or early April. In addition, the analysis includes controlling for a number of socioeconomic, demographic confounders, not adjusted for in other studies. Viral exposure is also adjusted for in the analysis. Use of small areas is also an advantage because air pollution varies considerably across large areas, such as local authority or region, used in other studies. The heterogeneity of pollution within a large area is considerable thus losing granularity. US counties are still relatively large, raising the question of how well such aggregated data can capture the local variation in confounding effects without being averaged out. The analysis in Netherlands used municipalities (Cole et al 2020). Although this is an improvement in terms of area size compared to the US county based analysis. Municipalities are also larger than MSOAs in England.

There are limitations to this study. Air pollution and covariates are all measured at the small area level not as individual level exposures, so like any ecological analysis there may be a danger of ecological fallacy. A few variables were not up to date but from the 2011 census. Future research that links individual data with pollution in residential areas should be carried out to avoid this issue. The study was observational and therefore any causal interpretation needs to be taken with caution. However, the causal pathways relating to respiratory and cardiovascular mortality has been established and it is highly likely that air pollution operates in the same way that impair the respiratory and circulation system which leads to exacerbates conditions of infected patients (Comunian et al, 2020).

## Conclusions

This analysis provides evidence that there is positive association between long term exposure to air pollution and COVID-19 deaths in England. Research on how modifiable factors may exacerbate COVID-19 symptoms and increase mortality risk is essential to guide policies and behaviours to minimize fatality related to the outbreak.

This paper provides further evidence that people living in an area with lasting high levels of pollutant are more prone to develop serious COVID-19 conditions. Future studies are needed to evaluate the role of the atmospheric pollution in certain populations and provide effective support for health intervention in reducing COVID-19 mortality. Reducing air pollution will have positive effect on many different areas reducing vulnerability to severe outcomes from virus infection like coronavirus. In addition, the intervention will have a lasting effect especially if the COVID-19 pandemic remains in the near future.

## Data Availability

Data will be available upon request

